# Social stressors associated with age-related T lymphocyte percentages in older US adults: Evidence from the Health and Retirement Study

**DOI:** 10.1101/2022.03.18.22272625

**Authors:** Eric T. Klopack, Eileen M. Crimmins, Steve W. Cole, Teresa E. Seeman, Judith E. Carroll

## Abstract

Exposure to stress is a well-established risk factor of poor health and accelerated aging. Immune aging, including declines in naive and increases in late memory and terminally differentiated T cells, plays an important role in immune health and tissue specific aging, and may contribute to the observed elevated risk for poor health among those who experience high psychosocial stress. However, past data have been limited in estimating the contribution of life stress to the development of accelerated immune aging and investigating mediators such as lifestyle and CMV infection, that might be useful points of intervention. The current study utilizes a national sample of 5744 US adults over the age of 50 to assess the relationship of social stress (viz., everyday discrimination, stressful life events, lifetime discrimination, life trauma, and chronic stress) with flow cytometric estimates of immune aging, including naive and terminally differentiated T cell percentages and the ratio of CD4^+^ to CD8^+^ T cells. Experiencing life trauma and chronic stress was related to a lower percentage of CD4^+^ naive T cells. Higher everyday discrimination, lifetime discrimination, and chronic stress were each associated with a greater percentage of terminally differentiated CD4^+^ T cells. Stressful life events, high lifetime discrimination, and life trauma were related to a lower percentage of CD8^+^ naive T cells. Stressful life events, high lifetime discrimination and chronic stress were associated with a higher percentage terminally differentiated CD8^+^ T cells. High lifetime discrimination and chronic stress was related to a lower CD4^+^:CD8^+^ ratio. Lifestyle factors and cytomegalovirus (CMV) seropositivity partially reduced these effects. Results identify psychosocial stress as a contributor to accelerating immune aging by decreasing naive and increasing senescent T cells.

## Introduction

Lifetime exposures to stressful conditions is a known risk factor for poorer health, increasing the risk for early onset of age-related disease and premature death(1–3). Models examining the mechanisms driving these effects have centered on the sequelae of repeated activation and prolonged activation of the sympathoadrenal and hypothalamic pituitary adrenal system, leading to wear and tear at the biological level (4). Many have postulated that this wear and tear manifests at the cellular level, causing accumulation of DNA damage, increasing inflammation, shortening telomere length and driving cellular aging (5, 6). Critically short telomere length within immune cells and cellular stress (e.g., DNA damage) can drive cells into a non-replicating state termed cellular senescence (7, 8). In addition to localized tissue-specific aging (9), age-related changes in immune function contribute to systemic aging, organ failure, and premature mortality, making immunosenescence a critical player in aging and disease (10).

Age is a robust determinant of immune cell population composition (11, 12), with aging immune systems characterized by a reduced pool of naïve B and T cells, increased pool of terminally differentiated T cells, increased supply of CD8^+^ and decreased supply of CD4^+^ T cells, and increased systemic inflammation (13). However, there is substantial variance in the rate of these changes and consequent alterations in immune cell composition among older adults (14). Understanding the contributors to this variance is critical given that immune aging is associated with chronic diseases (e.g., cancer (15) and cardiovascular disease (16)), weakened response to acute infections, increased risk of pneumonia, reduced efficacy of vaccines (17), and organ system aging (10).

Acute and chronic social stress can modify the immune system in several ways, including increased inflammatory signaling and reduced antiviral responses. (18, 19), suggesting that social stress may accelerate immune aging (20–30). However, past research has utilized small clinical and specialized samples, potentially limiting power to detect effects, power to assess mediators, and generalizability of results. In the present study, we utilized a nation-wide sample of 5641 US adults over the age of 50 to assess associations between four categories of social stress that have well established effects on heath (viz., everyday discrimination, lifetime discrimination, life trauma, and chronic stress) and enumerative measures of immunological aging, including naïve T cell percentages (CD3^+^/CD19^-^/CD45RA^+^/CCR7^+^/CD28^+^), terminally differentiated effector memory T cell percentages (TemRA: CD3^+^/CD19^-^/CD45RA^+^/CCR7^-^/CD28^-^) and the ratio of CD4^+^ to CD8^+^ T cells. Analyses also assessed potential mediation of these associations by socioeconomic and lifestyle factors and cytomegalovirus (CMV) seropositivity.

## Results

Descriptive statistics are shown in Table S1. The weighted sample is 55.2% female and has a median age of 67 years. 84.9% of the sample is Non-Hispanic White, 6.9% is Non-Hispanic Black, and 5.8% is Hispanic, and 2.5% is Non-Hispanic Other Race, 10.1% has less than 12 years of education, 30.7% has 12 years of education, 26.1% has 13-15 years of education, and 33.1% has 16 or more years of education. 9.4% of the sample are current smokers and 45.4% are former smokers, more than a third of the sample is obese (36.1%), and 53.6% were non-drinkers. On average, participants reported relatively low everyday discrimination, 0.50 stressful life events, 0.65 incidents of lifetime discrimination, 1.09 life traumas, and a moderate level of chronic stress. There was substantial variance on all of these scales.

Results from nested analyses regressing T cell subset percentage/ratio on each stressor while controlling for age, sex, race/ethnicity, and various potential mediators (education, drinking, smoking, BMI, and CMV serostatus), are shown in Figure 1. Individual figures and tables for each predictor are shown in Figures S1-5 and Tables S2-6.

**Figure 1.**
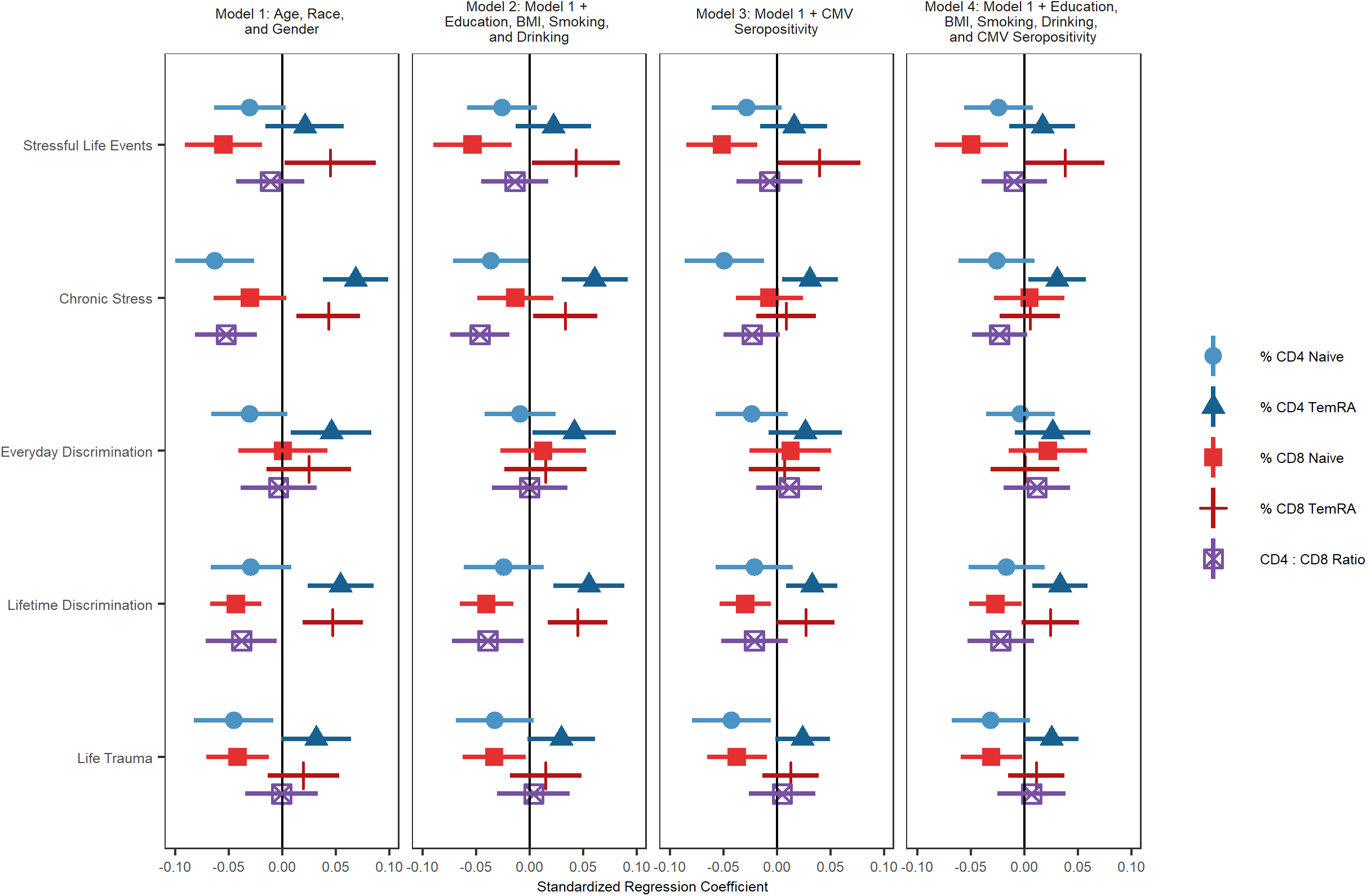
Regression coefficients and 95% confidence intervals from nested analyses regressing cell subset percentage/ratio on each stressor and mediators. All models control for age, race, and gender.

Experiencing more stressful life events was associated with a lower percentage of CD8^+^ naïve T cells and a higher percentage of terminally differentiated CD8^+^ T cells. The association between terminally differentiated CD8^+^ T cells and more stressful life events was reduced to non-significance after controlling for education, BMI, smoking, drinking and CMV seropositivity. The association between stressful life events and CD8^+^ naïve T cell percentage remained statistically significant after controlling for all covariates.

Experiencing greater chronic stress was related to a greater percentage of terminally differentiated CD4^+^ and CD8^+^ T cells, a lower percentage of CD4^+^ naïve T cells, and a lower ratio of CD4^+^ to CD8^+^ T cells. The association between chronic stress and CD4^+^ naïve T cells became non-significant after controlling for socioeconomic and lifestyle factors and CMV seropositivity. The associations between chronic stress and terminally differentiated CD8^+^ T cells and the CD4:CD8 ratio were reduced to non-significance after controlling for CMV seropositivity. The association between chronic stress and senescent CD4^+^ T cells remained significant even after adjusting for education, BMI, smoking, drinking, and CMV seropositivity.

Greater lifetime discrimination was related to having higher percentages of terminally differentiated CD4^+^ and CD8^+^ T cells, a lower percentage of CD8^+^ naïve T cells, and a lower CD4^+^:CD8^+^ ratio. The association between terminally differentiated CD8^+^ T cells and greater lifetime discrimination was reduced to non-significance after controlling for education, BMI, smoking, drinking and CMV seropositivity. The association between lifetime discrimination and the CD4^+^:CD4^+^ ratio was reduced to non-significance after controlling for CMV seropositivity. The associations between lifetime discrimination and terminally differentiated CD4^+^ and CD8^+^ naïve T cell percentages were both substantially reduced by these mediators but remained statistically significant.

Experiencing more everyday discrimination was related to having a higher percentage of terminally differentiated effector memory CD4^+^ T cells. This association was reduced after accounting for CMV seropositivity.

Finally, experiencing a greater number of life traumas was related to a smaller proportion of CD4^+^ and CD8^+^ naïve T cells. The association between CD4^+^ naïve T cells and trauma was reduced to non-significance after controlling for education, BMI, smoking, and drinking, suggesting that these socioeconomic and lifestyle factors may mediate this association. Associations between life trauma and CD8^+^ naïve T cells remained significant after including all covariates.

We also estimated models with interactions between stressors and gender and race/ethnicity, including all controls (not shown). There was only one statistically significant interaction. The effect of being experiencing more lifetime discrimination on naïve CD8^+^ T cell percentage was amplified for Hispanic respondents compared to Non-Hispanic White respondents.

## Discussion

Using a national sample of older US adults, we found that exposure to social stress was associated with T cell distributions indicative of accelerated immune aging. Specifically, life trauma and chronic stress were associated with a lower percentage of CD4^+^ naïve T cells, whereas everyday discrimination, lifetime discrimination, and chronic stress were associated with a greater percentage of terminally differentiated CD4^+^ T cells. Stressful life events, lifetime discrimination and life trauma were associated with a lower percentage of CD8^+^ naïve T cells, whereas stressful life events, lifetime discrimination, and chronic stress were significantly associated with higher percentage of terminally differentiated CD8^+^ T cells. Lifetime discrimination and chronic stress was associated with a lower CD4:CD8 ratio. These effects were all independent of chronological age, sex, and race/ethnicity.

All significant associations with CD4^+^ naïve T cells were reduced to non-significance after controlling for socioeconomic and lifestyle factors (viz., education, smoking, BMI, and alcohol use) and CMV seropositivity, suggesting that this may be an important pathway by which stress affects CD4^+^ T cell immunosenescence. This is consistent with past research suggesting that adiposity is associated with thymic involution (31) and research showing reduced percentages of CD4^+^ naïve T cells in people with alcohol use disorder (32). Further research is needed to clarify how these factors affect naïve CD4^+^ T cells.

Lifetime discrimination and chronic stress associations with terminally differentiated CD8^+^ T cells and the CD4:CD8 ratio became non-significant after controlling for CMV seropositivity and the association between stressful life events and terminally differentiated CD8^+^ T cells became non-significant after controlling for lifestyle and SES factors and CMV seropositivity. This is consistent with past research showing that stress is associated with impaired immunological control of latent viruses like CMV (19). More stressed individuals may have reduced control over and more frequent activation of CMV, leading to an increase in memory T cells and decrease in the CD4:CD8 ratio (33, 34).

The percentage of CD8^+^ naïve T cells remained significantly inversely associated with stressful life events, lifetime discrimination, and life trauma regardless of controls added to the model. This suggests that the accumulation of major negative events across the life course may have a direct effect on these naïve CD8^+^ T cells, although future research may uncover mediating mechanisms that were not identified here. This finding is again consistent with the possibility that thymic involution may be accelerated by social stress (e.g., via chronic activation of the HPA axis, which can accelerate thymic involution and thereby reducing naïve T cell development (18, 35)).

Finally, lifetime discrimination and chronic stress are associated with a greater percentage of terminally differentiated CD4^+^ T cells, regardless of controls in the model. CD4^+^ T cells play a critical role in cell-mediated immunity, producing cytokines that help direct the action of other cells. This compartment typically develops age related changes later than the CD8^+^ compartment (17); therefore, accelerated immune age phenotype in CD4^+^ T cells may be particularly indicative of aging immunity. Research has shown these senescent-like CD4^+^ T cells fail to optimally “help” B cell activation, produce more inflammatory cytokines, and have been implicated in cardiovascular disease (16). This cell subset may be of particular interest to researcher investigating stress and health (36).

The current study has several limitations. This is a study of community-dwelling older adults in the US. Future studies using non-US samples are needed. Our sample represents a cohort of older adults exposed to various forms of stress, discrimination, and trauma that might not be representative of younger cohorts or groups not well represented in HRS (e.g., indigenous peoples). We are not characterizing age-related changes across all cell types in the immune system. We selected T cells because of their importance in immune aging (see supporting information). Future research is needed to characterize stress effects on age-related changes in other cell types (e.g., B cells, monocytes, NK cells).

Despite these limitations, this study provides important insights on the role of social stress in immune aging, highlighting a key role for health behaviors and social-environmental conditions as correlates of naïve T cell decline as well as a distinctive association of stressors with higher terminally differentiated CD4^+^ T cell percentages (i.e., independent of variables controlled for here). These results raise the possibility that interventions such as CMV vaccination and senolytic therapies might potentially help reduce social disparities in T cell immunologic aging (13). Interventions aimed at reducing stress or increasing resilience may be needed to address these inequalities.

## Materials and Methods

### Sample

We utilize data from the Health and Retirement Study (HRS) 2016 Venous Blood Study (N = 9,934). Multiparameter flow cytometry was used to assess counts and percentages of 24 different types of immune cells using the standardized protocol by the Human Immunology Project (37) with minor modifications performed on an LSRII or a Fortessa X20 flow cytometer (BD Biosciences, San Diego, CA) More detailed methods for this sample are published elsewhere (38–40).

This sample was designed to be representative of the U.S. population when weighted. Stress scales were assessed as part of the psychosocial leave behind questionnaire at various waves. This questionnaire is given to a rotating random half of the core panel every other wave so that data is available for the full sample every four years. 200 participants were in a nursing home or were cohort ineligible in 2016, 614 participants were missing on one or more cell type, 3233 participants were missing on one or more stressor, and 142 participants were missing on one or more mediator or control variables. Thus, the final sample size is 5744 (see Figure S5).

### Measures

T lymphocyte distributions were assessed following the protocol described above. We focus here on the percentage of naïve CD4^+^ and CD8^+^ T cells (CD3^+^/CD19^-^/CD45RA^+^/CCR7^+^/CD28^+^) and terminally differentiated effector memory (TemRA) CD4^+^ and CD8^+^ T cells (CD3^+^/CD19^-^ /CD45RA^+^/CCR7^-^/CD28^-^). We also assess the ratio of CD4^+^ to CD8^+^ T cells using the quotient of counts of these compartments. Highly skewed cell variables (skewness > 2; viz., CD4^+^ TemRA and the CD4:CD8 ratio) were natural log transformed to approximate a normal distribution. These variables were standardized to have a mean of 0 and standard deviation of 1 to ease comparison.

We utilize five well-established health-relevant measures of social stress: stressful life events (41), chronic stress (42), everyday discrimination (43), lifetime discrimination (43), and life trauma (44). Because everyday discrimination, stressful life events, lifetime discrimination, and life trauma assess life course or ongoing exposure to stress, we use the first available data from the years these measures were assessed (2006-2016 for everyday discrimination, 2008-2012 for stressful life events, and 2006-2012 for lifetime discrimination and life trauma) Because we are interested in assessing ongoing chronic stress, we only use data from the 2014 and 2016 leave behind questionnaire. More information on these measures is available in the supporting information and more detailed information is available elsewhere (45).

Socioeconomic and lifestyle factors were assessed using self-reports of education (categorized: 0-11 years, 12 years, 13-15 years, and 16+ years as the reference), self-reported smoking (categorized: current smoker, past smoker, and non-smoker as the reference), BMI (categorized: ≥ 25 and < 30 overweight, ≥ 30 and < 35 obese 1, ≥ 35 obese 2, and < 25 normal and underweight as the reference), and alcohol use (categorized: 1-4 drinks per day drinking, 5+ drinks per day drinking, and non-drinker as the reference).

CMV seropositivity was assessed using IgG antibodies in serum with the Roche e411 immunoassay analyzer (Roche Diagnostics Corporation, Indianapolis, IN) (categorized: borderline, reactive, or non-reactive as the reference) (40).

In all models we control for chronological age, race/ethnicity (Non-Hispanic Black, Hispanic, Non-Hispanic other race, and Non-Hispanic White as reference), and gender (male as reference).

### Plan of Analysis

All stressors and T cell type percentages / ratios were standardized to have a mean of 0 and standard deviation of 1 to ease comparisons across models. A series of nested linear regressions were estimated regressing each T cell type percentage/ratio first on controls, then on controls and socioeconomic and lifestyle factors, then on controls and CMV seropositivity, and finally on controls, socioeconomic and lifestyle factors and CMV seropositivity. All analyses were conducted in R 4.1.1 (46) using the survey package (47).

## Data Availability

All data are publicly available: https://hrs.isr.umich.edu/about

## Acknowledgments

Support was provided by the USC/UCLA Center on Biodemography and Population Health through a grant from NIA (P30AG017265).

The HRS (Health and Retirement Study) is sponsored by the National Institute on Aging (grant number NIA U01AG009740) and is conducted by the University of Michigan.

## References

1. R. J. Turner, “Understanding health disparities: The promise of the stress process model” in Advances in the Conceptualization of the Stress Process, W. R. Avison, C. S. Aneshensel, S. Schieman, B. Wheaton, Eds. (Springer, 2009) https://doi.org/10.1007/978-1-4419-1021-9_1.

2. M. Kivimäki, A. Steptoe, Effects of stress on the development and progression of cardiovascular disease. Nat. Rev. Cardiol. 15, 215–229 (2018).

3. S. Cohen, “Psychological Stress, Immunity, and Physical Disease” in Scientists Making a Difference: The Greatest Livingbehavioral and Brain Scientists Talk about Their Most Important Contributions, S. Sternberg, S. Fiske, D. Foss, Eds. (Cambridge University Press, 2016).

4. B. S. McEwen, T. Seeman, Protective and Damaging Effects of Mediators of Stress: Elaborating and Testing the Concepts of Allostasis and Allostatic Load. Ann. N. Y. Acad. Sci. 896, 30–47 (1999).

5. E. S. Epel, et al., Accelerated telomere shortening in response to life stress. Proc. Natl. Acad. Sci. 101, 17312–17315 (2004).

6. I. Shalev, W. J. Hastings, “Psychosocial stress and telomere regulation” in Genes, Brain, and Emotions, (Oxford University Press, 2019), pp. 247–261.

7. L. C. Etzel, I. Shalev, “Effects of Psychological Stress on Telomeres as Genome Regulators” in Stress: Genetics, Epigenetics and Genomics, (Elsevier, 2021), pp. 109–117.

8. C. López-Otín, M. A. Blasco, L. Partridge, M. Serrano, G. Kroemer, The Hallmarks of Aging. Cell 153, 1194–1217 (2013).

9. B. G. Childs, M. Durik, D. J. Baker, J. M. van Deursen, Cellular senescence in aging and age-related disease: from mechanisms to therapy. Nat. Med. 21, 1424–1435 (2015).

10. M. J. Yousefzadeh, et al., An aged immune system drives senescence and ageing of solid organs. Nature 594, 100–105 (2021).

11. Z. Huang, et al., Effects of sex and aging on the immune cell landscape as assessed by single-cell transcriptomic analysis. Proc. Natl. Acad. Sci. 118 (2021).

12. B. Thyagarajan, et al., Age-related differences in T cell subsets in a nationally representative sample of people over age 55: Findings from the Health and Retirement Study. J. Gerontol. Ser. A (2021) https://doi.org/10.1093/gerona/glab300 (November 1, 2021).

13. A. Aiello, et al., Immunosenescence and Its Hallmarks: How to Oppose Aging Strategically? A Review of Potential Options for Therapeutic Intervention. Front. Immunol. 10 (2019).

14. K. J. Kaczorowski, et al., Continuous immunotypes describe human immune variation and predict diverse responses. Proc. Natl. Acad. Sci. 114, E6097–E6106 (2017).

15. W. X. Huff, J. H. Kwon, M. Henriquez, K. Fetcko, M. Dey, The Evolving Role of CD8+CD28−Immunosenescent T Cells in Cancer Immunology. Int. J. Mol. Sci. 20, 2810 (2019).

16. I. Broadley, A. Pera, G. Morrow, K. A. Davies, F. Kern, Expansions of Cytotoxic CD4+CD28− T Cells Drive Excess Cardiovascular Mortality in Rheumatoid Arthritis and Other Chronic Inflammatory Conditions and Are Triggered by CMV Infection. Front. Immunol. 8 (2017).

17. L. Pangrazzi, B. Weinberger, T cells, aging and senescence. Exp. Gerontol. 134, 110887 (2020).

18. R. Glaser, J. K. Kiecolt-Glaser, Stress-induced immune dysfunction: implications for health. Nat. Rev. Immunol. 5, 243–251 (2005).

19. J.-P. Gouin, L. Hantsoo, J. K. Kiecolt-Glaser, Immune Dysregulation and Chronic Stress among Older Adults: A Review. Neuroimmunomodulation 15, 251–259 (2008).

20. M. E. Bauer, Chronic Stress and Immunosenescence: A Review. Neuroimmunomodulation 15, 241–250 (2008).

21. A. Sommershof, et al., Substantial reduction of naïve and regulatory T cells following traumatic stress. Brain. Behav. Immun. 23, 1117–1124 (2009).

22. M. Maes, et al., The Effects of Psychological Stress on Leukocyte Subset Distribution in Humans: Evidence of Immune Activation. Neuropsychobiology 39, 1–9 (1999).

23. M. M. C. Elwenspoek, et al., T Cell Immunosenescence after Early Life Adversity: Association with Cytomegalovirus Infection. Front. Immunol. 8 (2017).

24. J. A. Bosch, J. E. Fischer, J. C. Fishcher, Psychologically adverse work conditions are associated with CD8+T cell differentiation indicative of immunesenescence. Brain. Behav. Immun. 23, 527–534 (2009).

25. A. E. Aiello, et al., Income and Markers of Immunological Cellular Aging. Psychosom. Med. 78, 657–666 (2016).

26. A. E. Aiello, et al., PTSD is associated with an increase in aged T cell phenotypes in adults living in Detroit. Psychoneuroendocrinology 67, 133–141 (2016).

27. K. E. Rentscher, et al., Chronic stress exposure and daily stress appraisals relate to biological aging marker p16INK4a. Psychoneuroendocrinology 102, 139–148 (2019).

28. C. P. Fagundes, R. Glaser, J. K. Kiecolt-Glaser, Stressful early life experiences and immune dysregulation across the lifespan. Brain. Behav. Immun. 27, 8–12 (2013).

29. J. K. Kiecolt-Glaser, L. McGuire, T. F. Robles, R. Glaser, Psychoneuroimmunology: Psychological influences on immune function and health. J. Consult. Clin. Psychol. 70, 537–547 (2002).

30. J. K. Kiecolt-Glaser, R. Glaser, Stress and immunity: Age enhances the risks. Curr. Dir. Psychol. Sci. 10, 18–21 (2001).

31. A. Kalathookunnel Antony, Z. Lian, H. Wu, T Cells in Adipose Tissue in Aging. Front. Immunol. 9, 2945 (2018).

32. P. Zuluaga, et al., Loss of naive T lymphocytes is associated with advanced liver fibrosis in alcohol use disorder. Drug Alcohol Depend. 213, 108046 (2020).

33. A. Bektas, S. H. Schurman, R. Sen, L. Ferrucci, Human T cell immunosenescence and inflammation in aging. J. Leukoc. Biol. 102, 977–988 (2017).

34. M. E. Bauer, M. D. la Fuente, The role of oxidative and inflammatory stress and persistent viral infections in immunosenescence. Mech. Ageing Dev. 158, 27–37 (2016).

35. M. E. Bauer, Stress, glucocorticoids and ageing of the immune system. Stress 8, 69–83 (2005).

36. L. I. Pearlin, The sociological study of stress. J. Health Soc. Behav. 30, 241–256 (1989).

37. H. T. Maecker, J. P. McCoy, R. Nussenblatt, Standardizing immunophenotyping for the Human Immunology Project. Nat. Rev. Immunol. 12, 191–200 (2012).

38. B. Thyagarajan, et al., Effect of delayed cell processing and cryopreservation on immunophenotyping in multicenter population studies. J. Immunol. Methods 463, 61–70 (2018).

39. H. Barcelo, J. Faul, E. Crimmins, B. Thyagarajan, A Practical Cryopreservation and Staining Protocol for Immunophenotyping in Population Studies. Curr. Protoc. Cytom. 84, e35 (2018).

40. E. M. Crimmins, J. D. Faul, B. Thyagarajan, D. R. Weir, “Venous blood collection and assay protocol in the 2016 Health and Retirement Study 2016 Venous Blood Study (VBS)” (2017).

41. R. J. Turner, B. Wheaton, D. A. Lloyd, The Epidemiology of Social Stress. Am. Sociol. Rev. 60, 104–125 (1995).

42. W. M. Troxel, K. A. Matthews, J. T. Bromberger, K. Sutton-Tyrrell, Chronic stress burden, discrimination, and subclinical carotid artery disease in African American and Caucasian women. Health Psychol. 22, 300–309 (2003).

43. D. R. Williams, Y. Yu, J. S. Jackson, N. B. Anderson, Racial Differences in Physical and Mental Health: Socio-economic Status, Stress and Discrimination. J. Health Psychol. 2, 335–351 (1997).

44. N. Krause, B. A. Shaw, J. Cairney, A Descriptive Epidemiology of Lifetime Trauma and the Physical Health Status of Older Adults. Psychol. Aging 19, 637–648 (2004).

45. Smith, Jacqui, Ryan, Lindsay H, Fisher, Gwenith G., Sonnega, Amanda, Weir, David R, “HRS Psychosocial and Lifestyle Questionnaire 2006-2016” (Survey Research Center, Institute for Social Research, University of Michigan, 2017).

46. R Core Team, R: A language and environment for statistical computing (R Foundation for Statistical Computing, 2021).

47. T. Lumley, Analysis of Complex Survey Samples. J. Stat. Softw. 9, 1–19 (2004).

48. P. A. Thoits, Stress and health: Major findings and policy implications. J. Health Soc. Behav. 51, S41–S53 (2010).

49. H. A. Turner, R. J. Turner, Understanding variations in exposure to social stress. Health (N. Y.) 9, 209–240 (2005).

50. T. P. Shippee, L. R. Wilkinson, M. H. Schafer, N. D. Shippee, Long-Term Effects of Age Discrimination on Mental Health: The Role of Perceived Financial Strain. J. Gerontol. Ser. B 74, 664–674 (2019).

51. D. Umberson, L. Hui, C. Reczek, Stress and health behaviour over the life course. Adv. Life Course Res. 13, 19–44 (2008).

52. E. M. Hailu, et al., Discrimination, social support, and telomere length: the Multi-Ethnic Study of Atherosclerosis (MESA). Ann. Epidemiol. 42, 58–63.e2 (2020).

