## Supporting Information for "Social stressors associated with age-related T lymphocyte percentages in older US adults: Evidence from the Health and Retirement Study"

*Why T cells?*

T cells are particularly affected in age-related immune changes. As a result of thymic involution, fewer naïve T cells are produced, and as a result of exposure to immunological insults, more memory T cells accumulate. CCR7 expression is depressed as central memory T cells become less common and effector memory T cells more common. CD45RA re-expression in effector memory T cells has been linked to cellular senescence. As part of a normal infection cycle, T cells reduce expression of CD28 (a critical costimulatory marker) to avoid over-activation after the infection subsides. After repeated cycles of activation, an abundance of $CD28^-$ T cells (especially $CD8^+$) accumulate. These cells are largely unable to respond to antigens, have limited reproductive ability, and generate inflammatory cytokines, potentially contributing to inflammaging (17). T lymphocytes may be of particular interest in light of the recent COVID-19 pandemic, given their important role in vaccine efficacy (17). Understanding variance in immunosenescence may be particularly important for understanding and combating age-related inequalities in vaccine efficacy and COVID-19 deaths.

*Why these stressors?*

Past theory and research on stress and health shows that there are three broad domains of health-relevant social stressors: acute negative events that resolve relatively quickly, chronic strains that represent ongoing difficulties, and traumas that represent major threats to a person's physical and mental wellbeing (48, 49). Research has demonstrated that these domains capture independent variance in health outcomes and are therefore all important to examine (48).

The five stressors addressed in the current study (everyday discrimination, stressful life events, lifetime discrimination, life trauma, and chronic stress) represent chronic stress related to discrimination, acute stressors from the prior five years, major acute stressors across the life course related to discrimination, life course exposure to major traumas, and ongoing chronic stressors, respectively. We thus address all three domains of stress identified in the literature. Additionally, the discrimination-relevant measures (everyday discrimination and lifetime discrimination) may be particularly relevant to the population of older US adults in the current study experiencing ageism and age-related discrimination (50).

Though there is early evidence that social stress accelerates immune aging (20, 24, 26, 27), past research has focused on individual stressors, limiting the conclusions that can be drawn about the effect of social stress on immune age phenotype, particularly with regard to the T cell compartment.

*Why these mediators?*

Past research shows that the immune system is highly affected by the socioeconomic and lifestyle factors identified here (18, 20). These health lifestyle factors are all also strongly related to chronic and acute stress and life traumas (51). Similarly, people who experienced a greater number of major traumas or acute stressful events across the life course may have obtained less education. Thus, these socioeconomic and lifestyle factors may explain why exposure to these stressors is associated with immune age phenotypic T cell distributions.

CMV seropositivity is particularly important for immune aging (23, 33). Past research suggests that repeated reactivation of CMV over the life course depletes naïve T cell reserves—which additionally diminish with age due to thymic involution—and increase the number of late differential T cells. Recent research shows that CMV can be activated by social stress via activation of the hypothalamic-pituitary-adrenal (HPA) axis and sympathetic nervous system

(SNS) (34). Thus, controlling for CMV may explain why chronic stress is associated with more immune age phenotypic T cell distributions.

*Stress Measures*

Stressful life events was assessed using a 6-item count of stressful life events that occurred in the past five years, including “have you involuntarily lost a job for reasons other than retirement”, “have you been unemployed and looking for work for longer than 3 months”, “was anyone else in your household unemployed and looking for work for longer than 3 months”, “have you moved to a worse residence or neighborhood”, “were you robbed or did you have your home burglarized”, and “have you been the victim of fraud” (41). The most common stressful life event was having someone looking for a job in your household (endorsed by about 13% of respondents) and the least common was moving to a worse residence (endorsed by about 3% of respondents).

Chronic stress was assessed using an 8-item scale both the number ongoing stressful problems and how distressing these problems are, including “health problems (in yourself)”, “physical or emotional problems (in spouse or child)”, “alcohol or drug use in family member”, “difficulties at work”, “financial strain”, “housing problems”, “problems in a close relationship”, and “helping at least one sick, limited, or frail family member or friend on a regular basis” with responses ranging from 1 (*no, didn’t happen*) to 4 (*yes, very upsetting*). Cronbach’s α for this scale is 0.64. Because chronic health problems could be confounded with immunosenescent leukocyte distribution, we also estimated the models using these same scales without that item. Results were highly similar with an identical pattern of significant results. We therefore use the full scale in all analyses. The most common chronic stressor was having an ongoing health problem (reported by about 67% of respondents) and the least common was ongoing housing problems (reported by about 17% of respondents).

Everyday discrimination was assessed using a 6-item scale focused on everyday hassles and ongoing chronic stress related to perceived discrimination (43). Items included “you are treated with less courtesy or respect than other people”, “you receive poorer service than other people at restaurants or stores”, “people act as if they think you are not smart”, “people act as if they are afraid of you”, “you are threatened or harassed”, and “you receive poorer service or treatment than other people from doctors or hospitals”, with responses ranging from 1 (*never*) to 6 (*almost every day*). Cronbach’s α for this scale is 0.80. The level of discrimination reported by respondents was highly similar to level reported in other studies (52). The most common form of discrimination reported was being treated with less courtesy or respect than other people (mean = 2.00) and the least common was receiving worse treatment from doctors or hospitals (mean = 1.23). Participants were also asked why they thought this happened to them. The most common first reason endorsed was age (endorsed by 1460 respondents), followed by gender (endorsed by 705 respondents), other reason (endorsed by 540 respondents), ancestry or national origin (endorsed by 424 respondents), race (endorsed by 319 respondents), weight (endorsed by 211 respondents), financial status (endorsed by 145 respondents), an aspect of their physical appearance (endorsed by 137 respondents), physical disability (endorsed by 92 respondents), religion (endorsed by 72 respondents), and sexual orientation (endorsed by 15 respondents).

Lifetime discrimination was assessed using a 7-item count of major stressful event related to discrimination throughout life, including “have you ever been unfairly dismissed from a job”, “for unfair reasons, have you ever not been hired for a job”, “have you ever been unfairly denied a promotion”, “have you ever been unfairly prevented from moving into a neighborhood because the landlord or a realtor refused to sell or rent you a house or apartment”, “have you ever been unfairly denied a bank loan”, “have you ever been unfairly stopped, searched, questioned, physically threatened or abused by the police”, and “have you ever been unfairly denied health care or treatment” (43). The most common form of discrimination was being dismissed from a job (endorsed by about 24% of respondents) and the least common was being prevented from moving to a neighborhood (endorsed by about 2% of respondents).

Life trauma was assessed using a count of 7 items focused on major traumatic events experienced throughout life, including “has a child of yours ever died”, “have you ever been in a major fire, flood, earthquake, or other natural disaster”, “have you ever fired a weapon in combat or been fired upon in combat”, “has your spouse, partner, or child ever been addicted to drugs or alcohol”, “were you the victim of a serious physical attack or assault in your life”, “did you ever have a life-threatening illness or accident”, and “did your spouse or a child of yours ever have a life-threatening illness or accident” (44). The most common trauma was having a spouse or child have a life-threatening illness (endorsed by about 25% of respondents) and the least common was firing a weapon or being fired upon in combat (endorsed by about 5% of respondents).

Table S1

Descriptive Statistics

| | Mean/ Proportion | SD | Range | | |
|---|---|---|---|---|---|
| CD4+ TemRA | -4.39 | 1.50 | -9.21 | - | -0.34 |
| CD4+ Naïve | 0.46 | 0.18 | 0.00 | - | 0.95 |
| CD8+ TemRA | 0.43 | 0.22 | 0.00 | - | 0.97 |
| CD8+ Naïve | 0.24 | 0.16 | 0.00 | - | 0.90 |
| CD4+:CD8+ Ratio | 1.15 | 0.71 | -2.37 | - | 3.68 |
| Stressful Life Events | 0.50 | 0.84 | 0.00 | - | 6.00 |
| Chronic Stress | 12.56 | 3.77 | 8.00 | - | 32.00 |
| Everyday Discrimination | 1.54 | 0.65 | 1.00 | - | 6.00 |
| Lifetime Discrimination | 0.65 | 1.00 | 0.00 | - | 7.00 |
| Life Trauma | 1.09 | 1.12 | 0.00 | - | 6.00 |
| Age | 68.34 | 9.23 | 50.00 | - | 107.00 |
| Gender (Female = 1) | 0.55 | | | | |
| Race | | | | | |
| White, not Hispanic | 0.85 | | | | |
| Black, not Hispanic | 0.07 | | | | |
| Hispanic | 0.06 | | | | |
| Other, not Hispanic | 0.02 | | | | |
| Education | | | | | |
| 16+ Years | 0.33 | | | | |
| 0-11 Years | 0.10 | | | | |
| 12 Years | 0.31 | | | | |
| 13-15 Years | 0.26 | | | | |
| BMI | | | | | |
| Under/Normal Weight | 0.27 | | | | |
| Overweight | 0.37 | | | | |
| Obese 1 | 0.22 | | | | |
| Obese 2 | 0.14 | | | | |
| Smoking | | | | | |
| Never Smoked | 0.45 | | | | |
| Current Smoker | 0.09 | | | | |
| Past Smoker | 0.45 | | | | |
| Alcohol Use | | | | | |
| Non-Drinker | 0.54 | | | | |
| 1-4 Drinks per day | 0.45 | | | | |
| 5+ Drinks per day | 0.02 | | | | |
| CMV Seropositivity | | | | | |
| Non-Reactive | 0.39 | | | | |
| Borderline | 0.02 | | | | |
| Reactive | 0.59 | | | | |

Note: SD = standard deviation; proportions may not sum to 1 due to rounding; T cell percentages / ratios and stressors are not standardized in this table; $CD4^+$ TemRA and the $CD4^+$:$CD8^+$ ratio are log transformed to approximate a normal distribution (see method section).

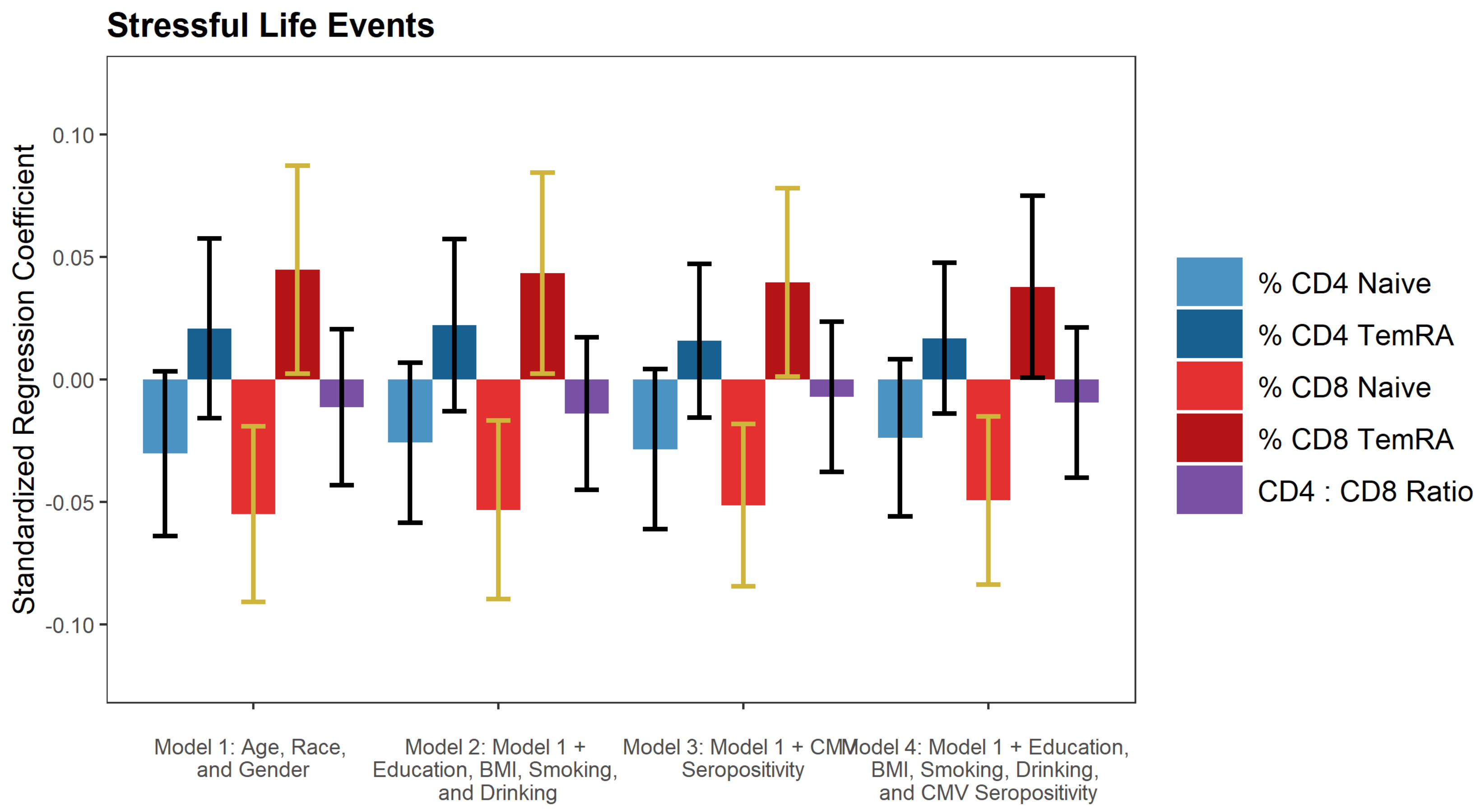


**Figure S1.** Results for Stressful Life Events

Note: gold confidence intervals indicate a statistically significant difference from 0.

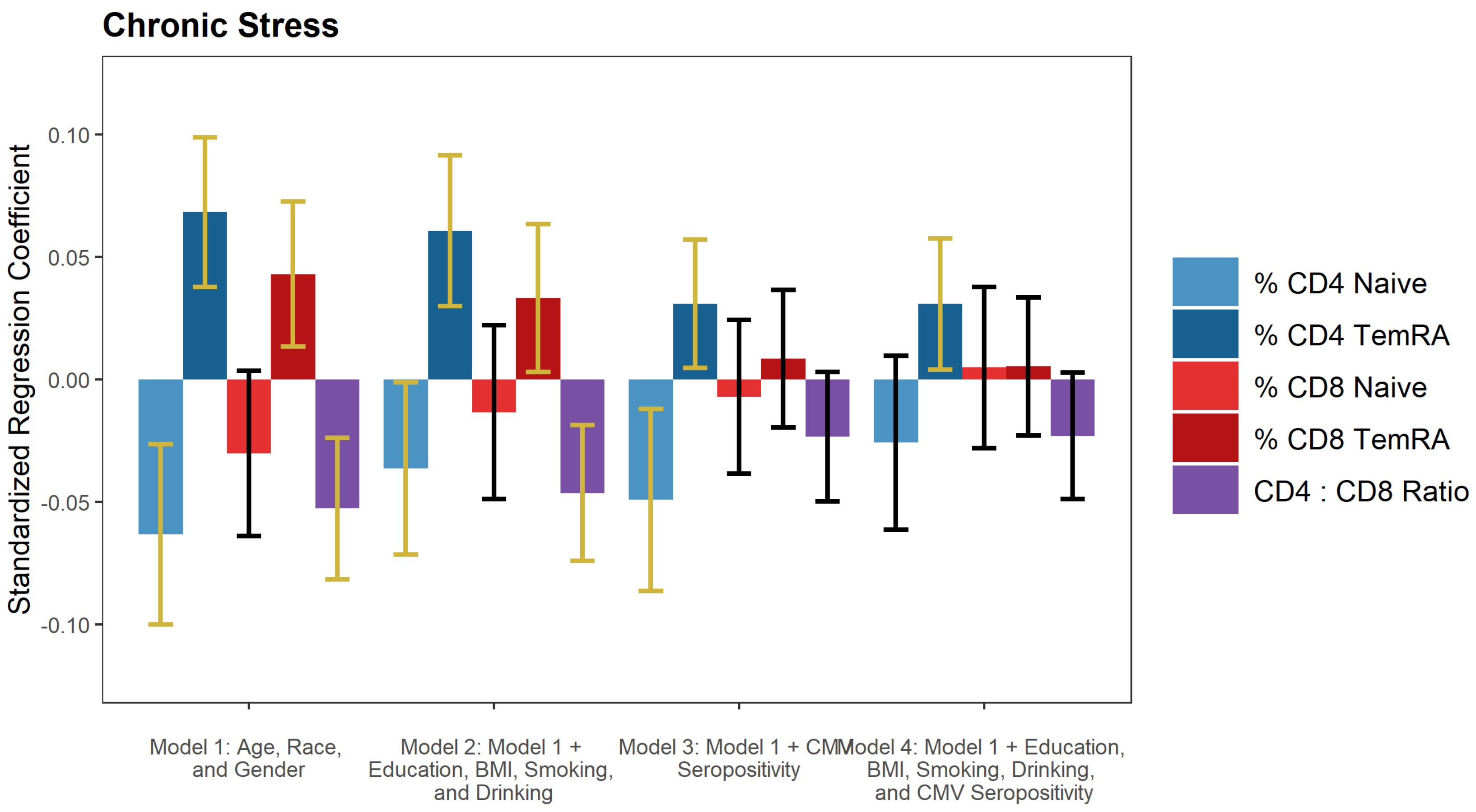


**Figure S2.** Results for Chronic Stress

Note: gold confidence intervals indicate a statistically significant difference from 0.

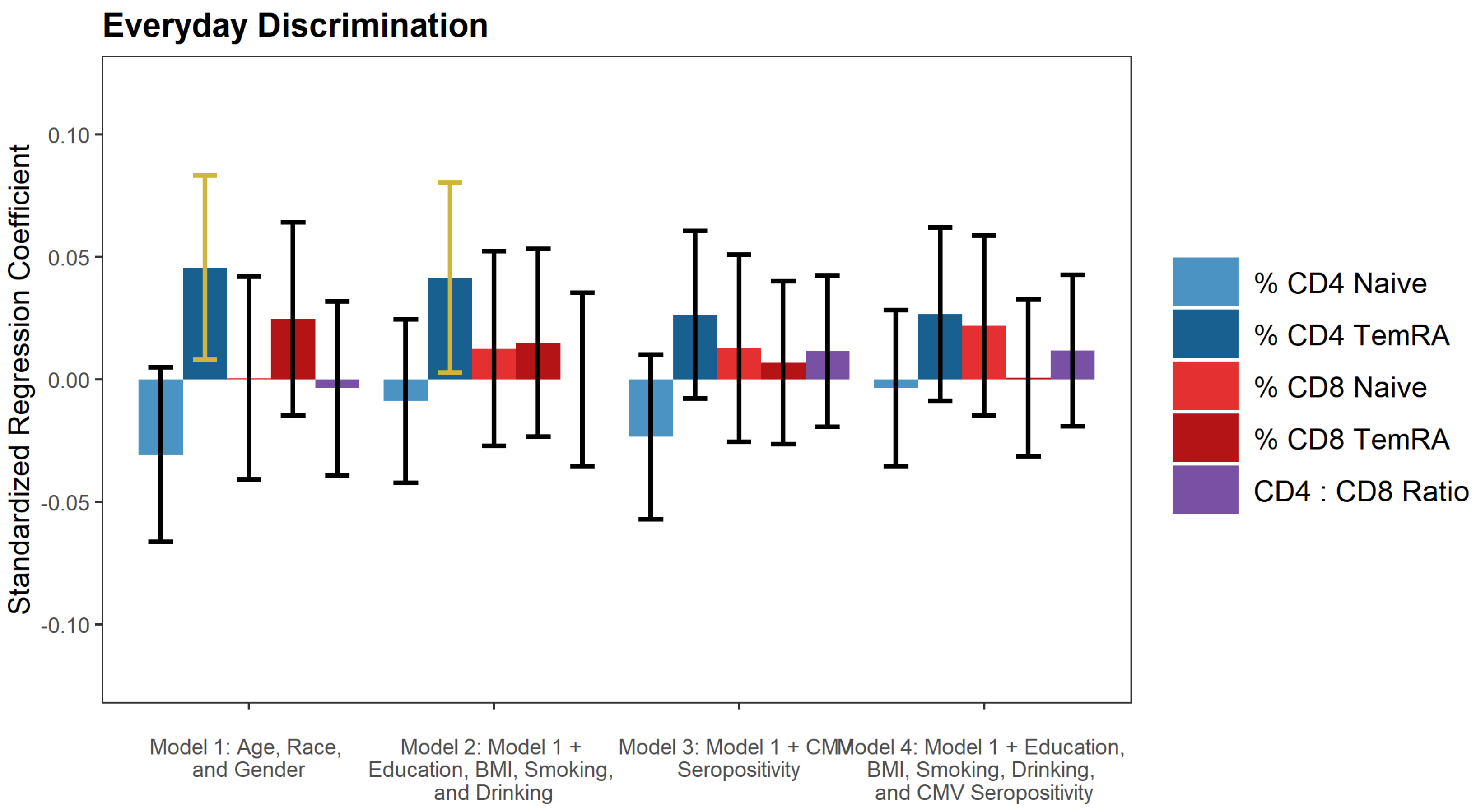


**Figure S3.** Results for Everyday Discrimination

Note: gold confidence intervals indicate a statistically significant difference from 0.

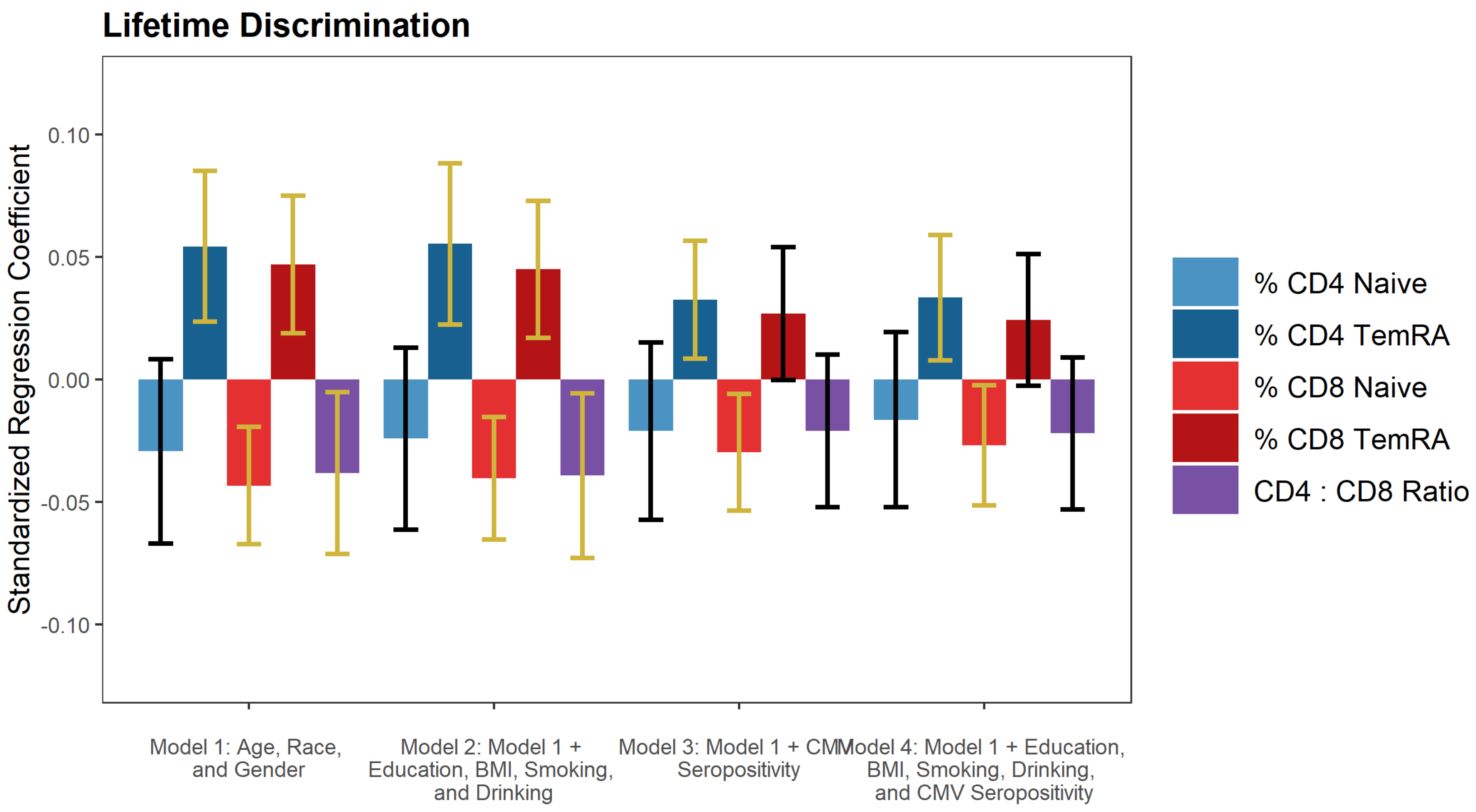


**Figure S4.** Results for Lifetime Discrimination

Note: gold confidence intervals indicate a statistically significant difference from 0.

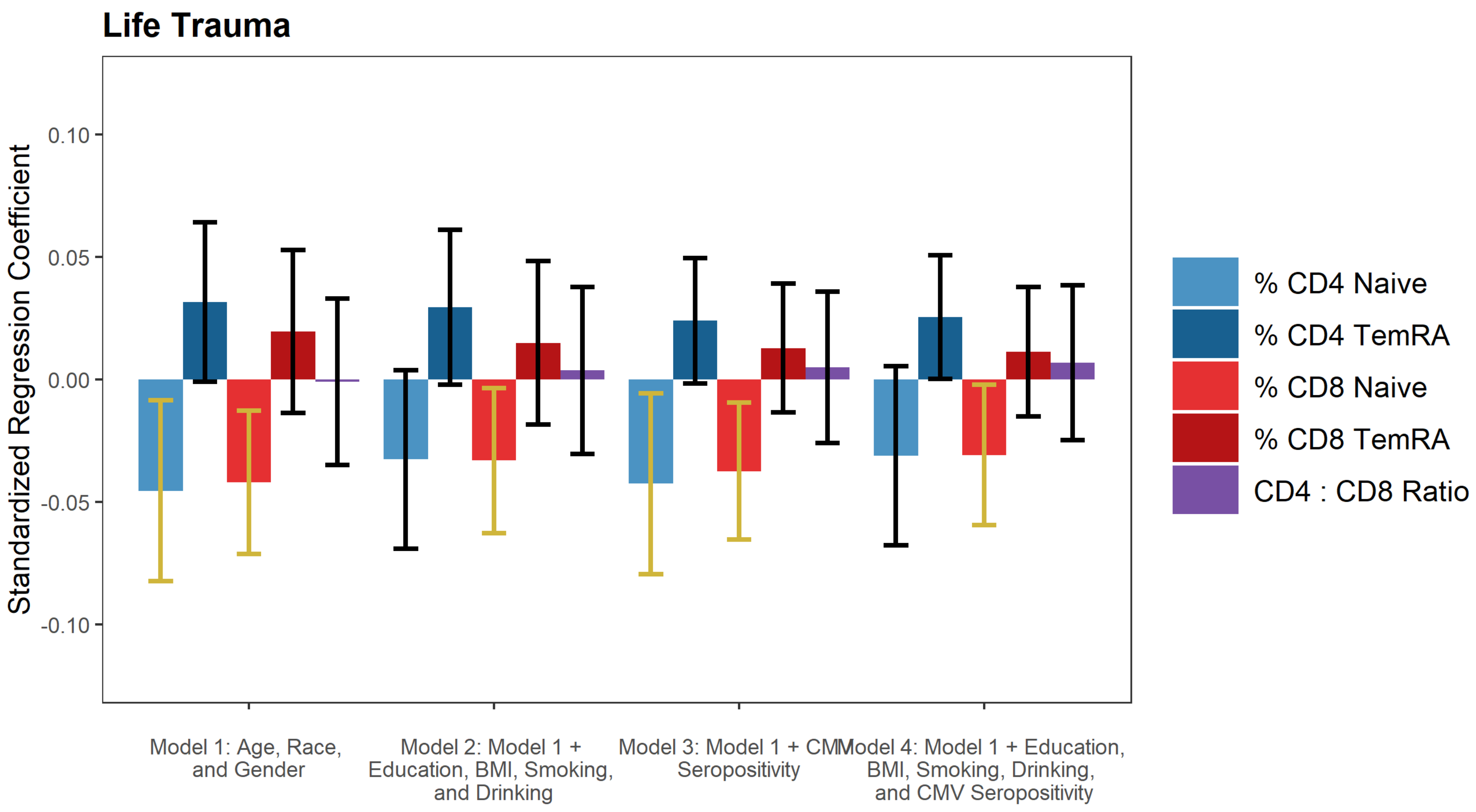


**Figure S5.** Results for Life Trauma

Note: gold confidence intervals indicate a statistically significant difference from 0.

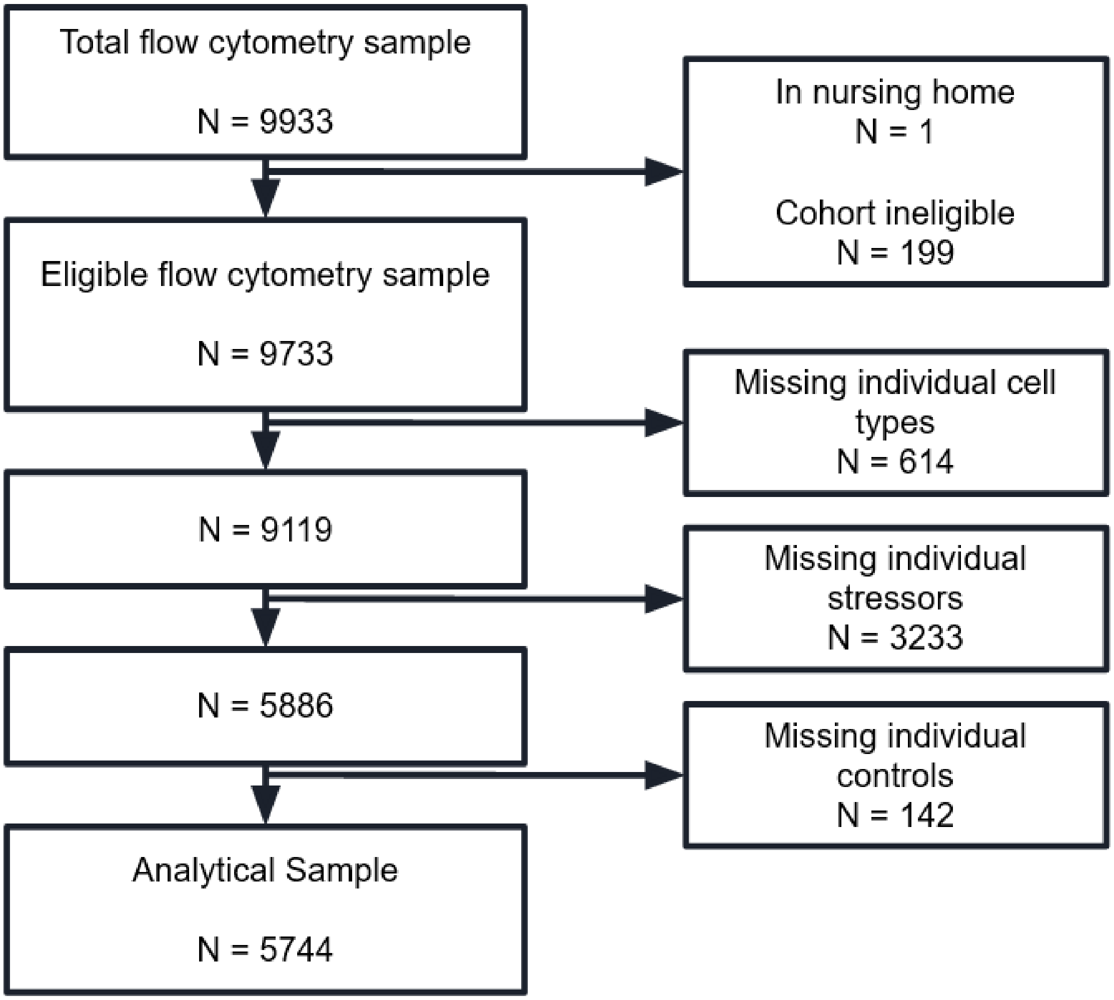


**Figure S6.** Sample Diagram
